# The cost-effectiveness of mammography-based female breast cancer screening in Canadian populations: a systematic review

**DOI:** 10.1101/2020.01.18.20018044

**Authors:** Talha Tahir, Melanie Mitsui Wong, Rabia Tahir, Michael Mitsui Wong

**Author notes:** Corresponding Author (TT). These authors contributed equally to this work. Address: McMaster University, 1280 Main Street West, Hamilton, Ontario, L8S 4L8.

## Abstract

**Introduction:** Mammography-based breast cancer screening is an important aspect of female breast cancer prevention within the Canadian healthcare system. The current literature on female breast cancer screening is largely focused on the health outcomes that result from screening. There is comparatively little data on the cost-effectiveness of the screening. Therefore, this paper sought to conduct a systematic review of the literature on the cost effectiveness of mammography-based breast cancer screening within female Canadian populations.

**Materials and methods:** A systematic review was performed in the PubMed database to identify all studies published within the last 10 years that addressed breast cancer screening and evaluate cost-effectiveness in a Canadian population.

**Results:** The search yielded five studies for inclusion, only three of which were applicable to average-risk Canadian women. The benefits of mortality reduction rose approximately linearly with costs, while costs were linearly dependent on the number of lifetime screens per woman. Moreover, triennial screening for average-risk women aged 50-69 years was found to be the most cost-effective in terms of cost per quality adjusted life year. The use of MRI in conjunction with mammography for women with the BRCA 1/2 mutation was found to be cost-effective while annual mammography-based screening for women with dense breasts was found to be cost-ineffective.

**Conclusion:** In spite of the growing interest to enhance breast cancer screening programs, analyses of the cost-effectiveness of mammography-based screening within Canadian populations are scarcely reported and have heterogeneous methodologies. The existing data suggests that Canada’s current breast cancer screening policy to screen average-risk women aged 50-74, biennially or triennially is cost-effective. These findings could be of interest to health policy makers when making decisions regarding resource allocation; however, further studies in this field are required in order to make stronger recommendations regarding cost-effectiveness.

## Introduction

Mammography-based female breast cancer screening is a modern staple of preventive healthcare [1]. It is administered in primary care facilities across Canada and the cost of the intervention is covered under Canada’s public healthcare system [1]. While there is no direct policy associated with screening for breast cancer, the government of Canada makes recommendations to primary care physicians and the general public about screening protocols [1]. It is currently recommended that women between the ages of 50-74 years who are at average-risk of developing breast cancer are screened via mammography biennially or triennially [1]. There are established benefits to breast cancer screening, particularly because of its high prevalence and stage variable mortality [2]. In Canada, breast cancer represents 25% of all cancer diagnoses among women and 13% of all cancer diagnoses overall [2]. However, as a result of the implementation of screening practices and improved therapeutic interventions, the mortality rate for female breast cancer has decreased by 44% since its peak in 1986 [2]. Since 2010, more than 80% of all female breast cancer diagnoses have occurred at stage I and II, with only 5% being diagnosed at stage IV [2]. However, the statistics on mortality rates and disease incidence only tell the story of breast cancer from an epidemiological perspective. To truly understand how to create effective policy for organized breast cancer screening across Canada, it is necessary to investigate the cost-effectiveness of current screening protocols.

Breast cancer screening recommendations are officially developed by the Canadian Task Force on Preventive Health Care (CTFPHC) [1]. The most recent breast cancer screening guidelines were released by the CTFPHC in 2018 [1]. While these guidelines and embedded recommendations incorporate a variety of factors, the emphasis is generally placed on patient costs and benefits. There is comparatively little exposure given to the financial costs of screening and false-positive scenarios. There is no analysis of cost-effectiveness within the current 2018 Canadian breast cancer screening guidelines and there is only a cursory discussion of the economic implications of screening within the older 2011 guidelines. Moreover, the two articles that were cited for cost-effectiveness in the 2011 CTFPHC guidelines were studies conducted in South Korea and the United States [3]. To make informed policy decisions about breast cancer screening, it is imperative to analyze Canadian data on cost-effectiveness [1,3]. Therefore, this systematic review intends to evaluate the cost-effectiveness of female breast cancer screening policies in Canadian populations through an analysis of current literature. The objective of this paper is to declare Canada’s current breast cancer screening policies as either cost-effective or ineffective and make recommendations that will increase cost-effectiveness in the future.

## Materials and methods

A systematic literature review for all original studies that addressed breast cancer screening and/or those that attempted to evaluate cost-effectiveness was conducted in accordance with the Preferred Reporting Items for Systematic Reviews and Meta-Analyses (PRISMA) Statement guidelines for reporting [4,5] (S1 Table. Criteria for literature search methods and inclusion criteria were determined a priori. No review protocol exists for this study. A systematic search was performed within the PubMed electronic database for articles published in peer-reviewed journals, with a full text available in English. Only papers published within the last 10 years were included to ensure the review was relevant to current breast cancer screening practices and treatment methods within Canada. In collaboration with an information specialist, we developed a search strategy tailored to the PubMed database using search terms such as “cost effectiveness”, “cost-benefit analysis”, “cost evaluation”, “Canada”, “screening”, “mammography”, “cancer”, “neoplasm”, “sarcoma”, and “breast” to identify studies investigating cost-effectiveness of mammogram screening in Canadian female populations (see S2 Table for the full search strategy).

### Eligibility Criteria

Title and abstract screening were conducted in duplicate by TT and MMW to identify studies for full-text review. The search was limited to studies that featured data on the cost-effectiveness of mammography-based female breast cancer screening policies within a Canadian demographic. Only English full-text, original research articles published within the past 10 years were eligible for inclusion. Any discrepancies raised during the screening process were resolved through consensus by TT and MMW. Full texts of selected publications were retrieved and assessed for eligibility by TT and MMW. Following full-text review, papers which contained original research related to breast cancer screening policies in Canada were included. Reference lists of included papers were also assessed to collect additional records and ensure that all relevant studies were captured.

### Data Extraction

Should the publication have met all eligibility criteria, the study characteristics, parameters of the model used, incremental cost-effectiveness ratios, average total cost of screening, incremental cost per death averted, and number of women screened to avert one death, were independently extracted by TT and MMW. If eligible studies did not report data necessary for inclusion in the systematic review, efforts were made to contact the corresponding authors directly.

### Risk of Bias & Quality Assessment

Risk of bias and study quality was assessed independently by TT and MMW using the Consolidated Health Economic Evaluation Reporting Standards (CHEERS) Checklist (S3 Table) [6]. This 24-item evaluation tool includes guidelines for critically appraising the quality of reporting economic model characteristics, analytic methods, study parameters, incremental costs and outcomes, amongst others [6]. For each item, evidence of its description within each study was reported. Final risk of bias evaluations were determined through consensus and review of source documents.

## Results

This search identified a total of 248 citations. After title and abstract screening, 29 articles were selected to undergo full text review. This initial round of screening did not apply the recency requirement of 10 years mentioned previously. Upon applying this recency requirement during the examination of full text articles, 5 papers were accepted and included in the systematic review (Fig 1). The characteristics of included articles are presented in Table 1. Please refer to S4 Table in the supplementary repository to view the articles excluded by full text screening and their reasons for exclusion.

**Table 1.** Characteristics and findings of included studies.

| Study Authors | Year | Findings |
| --- | --- | --- |
| Pataky et al. [7] | 2013 | Annual mammography plus MRI screening resulted in an ICER of \$50,900/QALY compared to annual mammogram screening alone. |
| Pataky et al. [8] | 2014 | Annual screening resulted in an additional 0.0014 QALYs compared to screening once every 2 years. |
| Gocgun et al. [9] | 2015 | Screening women aged 50-69 years once every 5 years is the most optimal with regards to cost per life saved. Annual mammogram screening is not cost-effective. |
| Mittmann et al. [10] | 2015 | Screening women aged 50-69 once every three years is the most cost-effective at \$83,070 per life year gained and \$94,762 per QALY. |
| Mittmann et al. [11] | 2018 | Annual screening per 1000 women costs the healthcare system \$7.4-10.7 million compared to a no-screening scenario, which would cost the system \$3.0 million per 1000 women. All screening programs increased life years gained and QALYs. |

**Fig 1.**
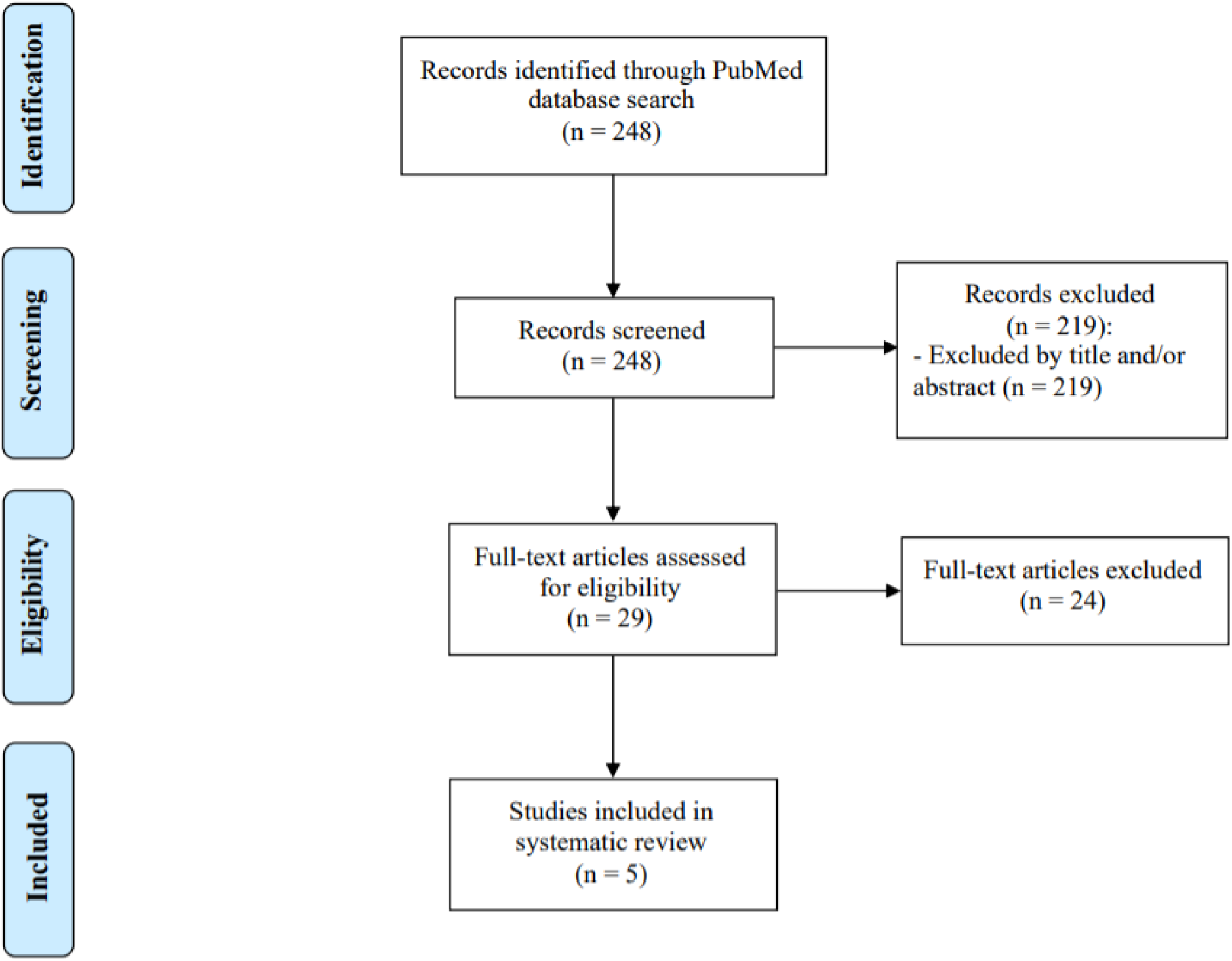
*PRISMA* diagram of literature review process for PubMed Search and Screening.

The Consolidated Health Economic Evaluation Reporting Standards (CHEERS) Checklist was applied to the 5 studies included in this review [6]. Upon evaluation, every article fulfilled most of the criteria on the checklist (S1 Table) and thus were deemed to be of good quality. All studies clearly presented well-defined research questions, characterized the study population, and provided justifications for the specific type of analytical model used. A notable difference between the articles was the presence of confidence intervals. The 2015 and 2018 papers by Mittman et al. did not contain confidence intervals or any other metric expressing uncertainty about their point estimates [10,11]. However, given that the papers incorporated a variety of cost, sensitivity, and incidence data to generate their models, this may explain the omission of confidence intervals within these studies.

This review takes a qualitative approach at evaluating the included studies. A meta-analysis could not be conducted due to the significant heterogeneity between the studies. In addition, the use of different models with varying input parameters, discounting rates, time horizons, and costs made it infeasible to statistically combine the studies.

The included studies varied in the populations studied, which were subdivided into two categories: those presenting a cost-effectiveness analysis for screening women at an average-risk for breast cancer, and those assessing cost-effectiveness for high-risk women. Three articles calculated the cost-effectiveness for screening average-risk women, Mittman et al 2015, Mittman et al 2018, and Gocgun et al 2015 (Table 2) [9–11].

**Table 2.** Characteristics of studies assessing cost-effectiveness for screening average-risk women.

| Study Authors (year) | Primary Outcome | Screening Intervention | Time Horizon | Costs |
| --- | --- | --- | --- | --- |
| Gocgun et al. (2015) [9] | Incremental cost per death averted | Film-based mammography | 20 years | 2015 Canadian dollars using Ontario-based costs for screening and treatment |
| Mittmann et al. (2015) [10] | Incremental cost per QALY | Digital mammography | Lifetime | 2012 Canadian dollars using Ontario-based costs for screening and treatment |
| Mittmann et al. (2018) [11] | Incremental cost per QALY | Digital mammography | Lifetime | 2012 Canadian dollars using Ontario-based costs for screening and treatment |

Both papers by Mittman et al. used the same University of Wisconsin Breast Cancer Epidemiology Simulation model. but addressed the topic from different perspectives [10,11]. The 2018 Mittman et al. paper considered cost-effectiveness from a single payer public healthcare system perspective, [11] whereas the 2015 paper by the same lead author considered cost-effectiveness from a societal perspective [10]. Both studies also used digital mammography as the screening intervention, expressed their costs in 2012 Canadian dollars using Ontario-based costs (Ontario Health Insurance Plan (OHIP)) for screening and treatment, and employed a lifetime horizon for cost calculation. The homogenous methodologies applied across these two papers thus made their respective estimates easy to compare.

The two Mittman papers concluded that the benefits of mortality reduction rose approximately linearly with costs, while costs were linearly dependent on the number of lifetime screens per woman. Then, the decision to screen is largely based on willingness to pay. To compare cost-effectiveness, an analysis of the incremental cost-effectiveness ratios (ICERs) of the two Mittman et al. studies was performed (Table 3).

**Table 3.** Cost-effectiveness of screening scenarios in Mittman et al. 2015 and Mittman et al. 2018. [10,11]

|  | **Triennial 50-69 years** | **Biennial 50-69 years** | **Annual 50-69 years** |
| --- | --- | --- | --- |
| **Mittman et al. 2015 [10]** | \$94,762/QALY | \$97,006/QALY | \$159,858/QALY |
| **Mittman et al. 2018 [11]** | \$36,918/QALY | \$38,142/QALY | \$65,944/QALY |

The 2015 Mittman et al. study had far higher ICERs due to its inclusion of costs to society, particularly loss in productivity [10]. Only 3 screening scenarios are examined in Table 2, as the 50-69 years age group closely approximates the current Canadian recommendation to screen average-risk women between 50-74 years of age. In both studies, the simulations with the 50-74 years age group were weakly dominated by the 50-69 years age group. Therefore, these studies provide support to the argument that average-risk women aged 70-74 years should not be screened using digital mammography.

The third study that examined cost-effectiveness for average-risk women was conducted by Gocgun et al. [9]. This study cannot be directly compared against the Mittman et al. studies because it did not calculate incremental cost per quality adjusted life year (QALY), instead opting to calculate incremental cost per death averted. Moreover, unlike the two Mittman et al. studies, Gocgun et al. did not use a lifetime horizon; instead, costs were evaluated over a horizon of 20 years. Gocgun et al. used data from the Canadian National Breast Cancer Screening Study to create and validate their model [9]. It should be noted that the data used to create this model was generated using film-based mammography, a screening technique that has since been replaced with digital mammography. The increased sensitivity of digital mammography may have resulted in more cost-effective outcomes. Ontario (OHIP) data expressed in 2015 Canadian dollars was used to input costs of screening and treatment scenarios [9].

Gocgun et al. (2015) concluded that more aggressive screening scenarios generate greater health benefits but at an increased cost (Table 4) [9]. Screening average-risk women aged 50-69 years every 5 years was found to have the lowest incremental costs, but the highest number of women screened to avert one death. Biennial screening for average-risk women between 50-69 years had the most favorable combination of incremental cost per death averted and number of women screened to avert one death. Similar to Mittman et al., Gocgun et al. found a sharp increase in cost per utility, or in this case, cost per death averted when comparing biennial or triennial screening to annual screening [9].

**Table 4.** Cost-effectiveness of select screening scenarios from Gocgun et al. 2015. [9]

|  | **Triennial 50-69 years** | **Biennial 50-69 years** | **Annual 50-69 years** |
| --- | --- | --- | --- |
| **Average total cost (screening, diagnosis and treatment costs)** | \$16,298,486 | \$18,414,393 | \$24,091,226 |
| **Incremental cost per death averted** | \$626,973 | \$654,940 | \$981,265 |
| **Number of women screened to avert one death** | 1,332 | 960 | 785 |

The remaining 2013 and 2014 studies by Pataky et al. included in this review focused on populations at high-risk for breast cancer (Table 5) [7,8]. Both studies constructed their Markovian model based on breast cancer incidence, treatment, and screening costs in British Columbia using a lifetime horizon for the calculation of costs. The 2013 study by Pataky et al. examined cost-effectiveness of using Magnetic Resonance Imaging (MRI) in conjunction with mammography for women carrying the BRCA1/2 mutation, expressing costs in 2008 Canadian dollars [7]. Women with the BRCA1/2 mutation are at a higher risk of developing breast cancer, therefore, they may benefit from the higher sensitivity of the MRI [12]. Pataky et al. (2013) found an ICER of $50,911/QALY for using MRI in conjunction with mammography instead of using mammography alone [7]. At a willingness to pay of $100,000/QALY, the use of MRI screening is cost-effective 85.6% of the time. However, Pataky et al. found the model to be highly dependent on the cost of an MRI scan, which may change the applicability of the result in locations where an MRI scan is more expensive [7].

The 2014 study by Pataky et al. investigated the cost-effectiveness of annual vs. biennial mammography screening for women with dense breasts, expressing costs in 2007 Canadian dollars [8]. The sensitivity of mammography screening was significantly lower in women with dense breasts, consequently making them more likely to be diagnosed with interval cancers [13]. The study found that screening breast cancer in women with dense breasts annually instead of biennially has an ICER of $565,912/QALY [8]. The extremely high ICER coupled with the fact that only 43.4% of the simulations showed annual screening to reduce QALYs relative to biennial screening suggest the cost-ineffectiveness of this strategy [8]. Moreover, the sensitivity data used in this study was for film-based mammography instead of digital mammography, making it difficult to judge cost-effectiveness in a healthcare environment dominated by digital mammography. Based on this study, annual breast cancer screening for women with dense breasts can effectively be considered cost-ineffective until further research proves otherwise [8].

**Table 2.** Characteristics of studies assessing cost-effectiveness for screening high-risk women

| Study Authors (year) | Primary Outcome | Screening Intervention | Time Horizon | Costs |
| --- | --- | --- | --- | --- |
| Pataky et al. (2013) [7] | Incremental cost per QALY | Magnetic resonance imaging in conjunction with mammography | Lifetime | 2008 Canadian dollars using British Columbia-based costs for screening and treatment |
| Pataky et al. (2014) [8] | Incremental cost per QALY | Film-based mammography | Lifetime | 2007 Canadian dollars using British Columbia-based costs for screening and treatment |

## Discussion

To the best of our knowledge, this is the first systematic review that assesses the cost effectiveness of mammography-based breast cancer screening within Canadian female populations. The results of the 5 studies included in this review ultimately suggest that aggressive screening strategies incur higher costs, but they also provide more health benefits. The optimal strategy for any given situation is decided largely by willingness to pay. However, a public healthcare system like Canada must make concrete decisions on which screening practices to implement based on clinical and cost-effectiveness. To make such a decision, it is necessary to understand the thresholds for cost-effectiveness within the field of oncology.

According to a 2010 paper that evaluated the attitudes of US and Canadian physicians on cost-effectiveness, the majority of oncologists in both Canada and the United States see $100,000/life-year as the threshold for cost-effective oncology treatments [14]. Traditionally, when examining healthcare interventions, the threshold of $50,000/QALY (USD) has been cited [15]. However, in recent literature, the trend has been to use a threshold of $100,000/QALY with the World Health Organization and many economists arguing that thresholds of anywhere from $110,000 to $160,000 per QALY should be used in a population like the United States, given median household income and other factors [15]. If one were to extrapolate these results to Canada, it seems that at the very minimum a value of $100,000/QALY should be used when evaluating the cost-effectiveness of breast cancer screening procedures.

Having made this assumption, we find that the estimates given by both the 2018 and 2015 Mittman et al. studies for biennial and triennial screening of average-risk women between the ages of 50-69 years fall below the threshold of $100,000/QALY [10,11]. It seems that even with the inclusion of societal costs such as loss in productivity, biennial and triennial screening of women aged 50-69 years is cost-effective. The results of the study by Gocgun et al. cannot be analyzed by the metric of cost-effectiveness we have used above, as the study did not integrate QALYs in its incremental cost-effectiveness ratios. Moreover, Gocgun et al.’s study, more than any other in this review, suffered from a poorly constructed model that was forced to make multiple simplifying assumptions due to data scarcity. For example, due to data scarcity, Gocgun et al. assumed the stage distribution of clinically presenting cancer for the 60-69 age group to be the same as that for the 50-59 age groups [9]. As a result, we will exclude Gocgun et al. results in making a decision about the cost-effectiveness of Canada’s current breast cancer screening policies.

The 2014 study by Pataky et al. on the cost-effectiveness of annual vs. biennial mammography breast cancer screening for women with dense breasts found an ICER of $565,912/QALY [8]. The high ICER and the variation seen in the simulations for annual versus biennial breast cancer screening for women with dense breasts leaves us uncertain about its cost-effectiveness. Lastly, the 2013 Pataky et al. study looking at using MRI screening in conjunction with mammography on women carrying the BRCA1/2 mutation found an ICER of $50,911/QALY as compared to mammography screening alone [7]. This is well below our cost-effectiveness threshold of $100,000/QALY and suggests that using MRI in conjunction with mammography is indeed cost-effective for that specific population of women.

The consensus seems to be that the current breast cancer screening policies of Canada, to screen average-risk women aged 50-74 years biennially or triennially, are cost-effective. However, the results of the above studies suggest that it is cost-ineffective to screen women with dense breasts annually and that it is cost-effective for women with the BRCA1/2 mutation to be given the option of using MRI in conjunction with digital mammography.

The results of the review must be tempered by the heterogeneity and limitations of the constitutive studies. The included papers used different models that were validated with different reference data, utilized different time horizons, and conducted different numbers of simulations. Moreover, the cost-effectiveness of breast cancer screening programs tends to be highly sensitive to the discounting rate, as many of the costs are accrued near the beginning of the program while the benefits are reaped near the end. Every study used a different discounting rate with 1.5% as the lowest and 5% as the highest. The methodological differences between studies mean that the results of this review must be interpreted with caution and that it is difficult to make firm statements about the cost-effectiveness of breast cancer screening in Canada.

Despite the heavy focus on optimizing breast cancer screening protocols, there is a scarcity of literature available on the cost-effectiveness of breast cancer screening within Canadian populations. Further studies are required to be able to make stronger recommendations regarding cost-effectiveness, which will prove critical in making decisions regarding resource allocation. The concerns presented in this review could serve as a basis for consideration by health policy makers, who must consider cost as a major factor when evaluating mammogram screening programs.

## Data Availability

Relevant data have been included in the supplemental figures. Any additional data can be obtained upon request by emailing

## Supporting information

**S1 Table. PRISMA Checklist. S2 Table. Search strategy**.

**S3 Table. CHEERS Checklist for Included Articles**.

**S4 Table. Papers excluded from full text review along with reasons for exclusion**.

